# Impact of HPV vaccination in Spain: Reductions in vaccine-type genotypes, cytological abnormalities, and the influence of epidemiological factors

**DOI:** 10.64898/2026.09.10.26362440

**Authors:** Alicia Inés García Señán, Edurne Arenaza Lamo, Antonia Dávila Expósito, Maialen Arruti Martín, Jhon William Comba Miranda, David del Valle Peña, María Raquel Ramos Triviño, Sara Palomo Cousido, Beatriz Dominguez Eguizabal, Maite Cortés Antolín, María José Puente Martínez, Tamara Zudaire Fuertes, Jesus Javier Sola Gallego, Carolina Ruiz Sánchez, Horacio Gil, Spanish HPV Vaccine Impact Study Group

## Abstract

Data evaluating human papillomavirus (HPV) vaccine impact in Spain remain limited. We aimed to assess HPV and cytology findings among vaccinated and unvaccinated women as an early indicator of its effectiveness. During April 2024-January 2026, a total of 908 women participating in a population-based cervical cancer screening in three regions of Spain were recruited (58% vaccinated). Cervical samples and epidemiological and vaccination data were obtained. HPV was detected in 37% (337/908) and high-risk HPV in 24% (221/908). High-grade cytological lesions (1.2%, 11/907) were more frequent in unvaccinated women (2.4% vs 0.38%, p=0.011) and showed higher prevalence of HPV 16/18 (5.5% vs 0.76%; aOR: 4.9; 95% CI: 1.4–17) and HPV31 (3.4% vs 1.3%; aOR: 2.7; 95% CI: 1.0–6.9) compared with vaccinated women. HPV51 and HPV70 were more frequent among vaccinated women; however, both were associated with factors unrelated to vaccination. HPV51 was linked to a higher number of sexual partners (aOR: 2.6; 95% CI: 1.2–5.6), while HPV70 to smoking (aOR: 0.36; 95% CI: 0.13–0.99). Lower detection of HPV 16/18, HPV31 and high-grade lesions among vaccinated women supports vaccine effectiveness and cross-protection, highlighting the importance of continuous comprehensive surveillance of HPV genotype, including epidemiological and sexual behavioral factors.

## INTRODUCTION

Human papillomavirus (HPV) is the primary causative agent of cervical cancer and is also associated with other anogenital and head and neck cancers; recurrent respiratory papillomatosis; and anogenital warts^1,2^. Despite the availability of effective preventive measures, cervical cancer remains a major public health challenge worldwide, particularly in low- and middle-income countries, with an estimated 604,000 new cases and 280,000 deaths reported in 2024^3^.

In 2020, the World Health Organization (WHO) launched a global strategy for the elimination of cervical cancer, based on achieving the 90–70–90 targets: 90% of girls fully vaccinated against HPV by 15 years of age, 70% of women screened by 35 and 45 years, and 90% of women with cervical disease receiving appropriate treatment. This comprehensive approach aims to reduce cervical cancer incidence below 4 cases per 100,000 women-years worldwide by 2030^4^.

Six WHO-prequalified HPV vaccines are available^5^: bivalent vaccines (2vHPV) targeting HPV 16 and HPV 18, quadrivalent vaccines (4vHPV) that additionally protect against non-oncogenic HPV 6 and HPV 11, and a nonavalent vaccine (9vHPV) that further covers five additional genotypes, providing protection against HPV genotypes responsible for 77% to 95% of cervical cancers^6^.

In Spain, cervical cancer remains an important public health issue, with an annual incidence rate of 8.2 cases per 100,000 women^7^. Routine HPV vaccination for girls aged 11–14 years was introduced in 2007–2008 as a publicly funded immunization program, using either the 2vHPV or 4vHPV, and was progressively expanded to include catch-up programs, high-risk groups, and adolescent boys from 2022 onwards^8^. Nowadays, vaccine coverage reached 91% (one dose) and 86% (two doses) among 15-year-old girls in 2024^9^. In parallel, a population-based cervical cancer screening program was approved in 2019 and has been gradually implemented across the Spanish regions^10^.

Although HPV vaccination and screening programs have been implemented in Spain, data assessing their population-level impact remain limited^11-15^, and no nationwide HPV molecular surveillance system has yet been established to monitor changes in HPV genotype distribution over time.

The aim of this study is to characterize the HPV genotype distribution and cytological abnormalities in women participating in the population-based cervical cancer screening program, assessing the impact of HPV vaccination while considering relevant epidemiological and behavioral factors.

## METHODS

### Study design and patient selection

A cross-sectional study was conducted between April 2024 and January 2026. Women born between 1992 and 1997 who participated in the population-based cervical cancer screening in three regions of Spain— Guadalajara (total female population (TFP): 143,390), La Rioja (TFP: 165,880) and the Basque Country (TFP: 1,150,802)^16^—were invited to participate in the study. This cohort includes the first vaccinated women (VW) according to the national immunization program introduced in Spain in late 2007, as well as a substantial proportion of unvaccinated women (UVW) within a relatively narrow birth cohort. 4vHPV was used at that time for the immunization campaign in these regions. Women without a cervix (i.e. prior hysterectomy or cervical conization) and those who had not initiated sexual activity were excluded from the study. After providing written informed consent, an online questionnaire with epidemiological, clinical, HPV vaccination and sexual-behavior data of the participants was completed by a midwife interviewing the participant and with the available vaccination records (Supplementary material) and labeled with a non-traceable study code to ensure irreversible anonymization. This study was approved by the Committee for Ethics in Research with Medicine of La Rioja (CEImLAR Eom 141), Guadalajara (CEIm 2024.05.PR) and Euskadi (CEIm-E PI2024044).

### Cytology testing and HPV genotyping

As part of the screening program routine, cervical brush samples were collected from the participants for liquid-based cytology (PreservCyt; Hologic Inc., MA, USA). Cytological analysis was performed using the ThinPrep Pap Test (Hologic Inc.) by the reference screening laboratory in each region. Results were reported according to the 2014 Bethesda classification^17^. After completion of routine screening analyses, the cytology result and an aliquot of each sample was labeled with the same non-traceable code used in the questionnaire and sent to the National Center for Microbiology-Instituto de Salud Carlos III (CNM-ISCIII) for complete HPV genotyping.

Prior to HPV detection testing, DNA was extracted from 1 ml of the sample using the MagNA Pure 24 Total Nucleic Acid Kit (Roche Life Science). HPV genotyping was performed using the Allplex HPV28 Detection Assay (Seegene, South Korea), a multiplex real-time PCR assay that independently detects 13 high-risk genotypes (HR-HPV) (16, 18, 31, 33, 35, 39, 45, 51, 52, 56, 58, 59, and 68) and 15 low-risk genotypes (6, 11, 26, 40, 42, 43, 44, 53, 54, 61, 66, 69, 70, 73, and 82), in addition to the human β-globin gene, which is used as an internal control.

In cases where any HR-HPV, HPV 6, or HPV 11 was detected, the viral load (VL) was estimated using serial dilutions of reference plasmids for each HR-HPV type, HPV 6, and HPV11, obtained from the International HPV Reference Center (Karolinska Institute, Sweden), except for HPV 39 and the human β-globin gene, which targets amplified by the Allplex HPV28 Detection Assay were cloned using TOPO TA Cloning® (Invitrogen, Carlsbad, CA, USA), following the manufacturer’s instructions. VL was expressed as copies per 1,000 human cells.

Women who participated in the study followed the established screening algorithms according to the results obtained in the regional reference laboratories. At the time of recruitment, cytology was the primary cervical cancer screening test for women under 35 years of age, with HPV testing performed only in cases of abnormal cytology^10^. Genotype results obtained in the CNM-ISCIII were used exclusively for research purposes, thus avoiding overdiagnosis and interference with the established screening algorithms.

### Data analysis

VW were defined as participants who received at least one dose of any type of vaccine. Full vaccination was considered when two or three doses were administered according to the age recommendations. To evaluate the association between categorical variables and vaccination status, Chi-square or Fisher’s exact tests were used, as appropriate.

Continuous variables were analyzed using Student’s t-test or the Mann–Whitney U test, depending on data distribution. Statistical significance was defined as a p-value < 0.05.

Associations between the variables defined in the study, and the detection of the different HPV genotypes, were estimated using odds ratios (ORs) with their corresponding 95% Confidence Intervals (95% CI). Subsequently, a multivariable logistic regression model was constructed, which included those variables that were statistically significant in the univariable analysis. All statistical analyses were performed using STATA statistical software, version 18 (StataCorp, College Station, TX, USA).

## RESULTS

### Participants and collected data

A total of 908 women from the Basque Country (n = 410; 45%), Guadalajara (n = 302; 33%) and La Rioja (n = 196; 22%) were enrolled in the study between April 2024 and January 2026 (Table 1). Overall, 58% (527/908) of participants had received at least one dose of an HPV vaccine. Notably, VW and UVW differed in several characteristics (Table 1). UVW showed significant differences compared with VW regarding origin, marital status, parity and age (p < 0.001), including a higher proportion of women from Latin America (46% vs 10%), a higher proportion of married women (43% vs 16%), higher parity (≥2 births: 18% vs 4.0%) and a higher mean age [30.1 Standard Deviation (SD)± 1.7 vs 28.9 SD± 1.5 years]. In addition, statistically significant differences were observed in the number of sexual partners (p = 0.011) and condom use (p < 0.001), with UVW reporting fewer sexual partners (1–5 partners: 66% vs 59%) and a higher frequency of no condom use in the previous year (43% vs 34%) (Table 1).

**Table 1:**
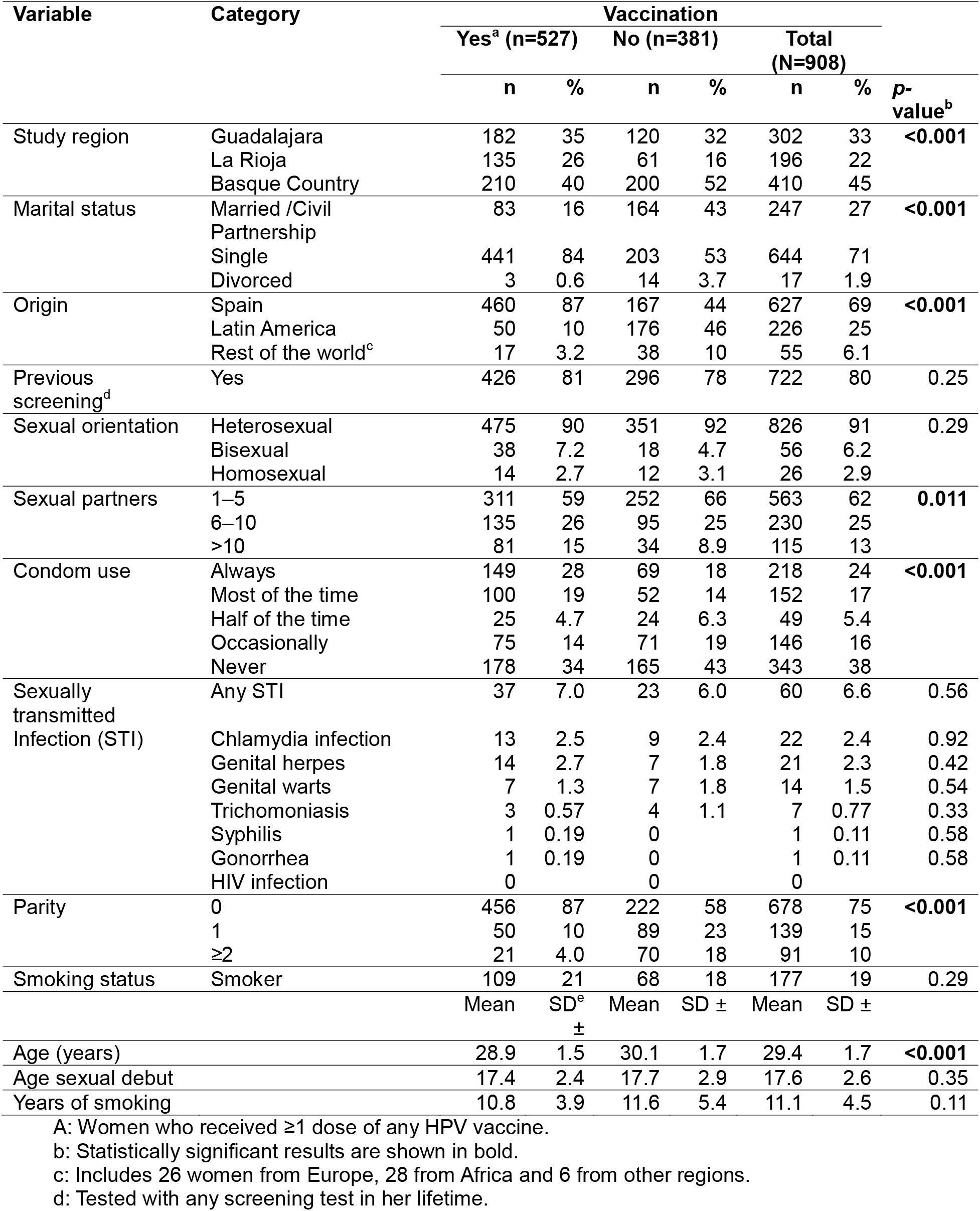
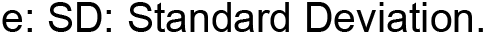
Characteristics of the study population according to the vaccination status.

| Variable | Category | Vaccination |  |  |  |  |  | p-value<sup>b</sup> |
| --- | --- | --- | --- | --- | --- | --- | --- | --- |
|  |  | Yes<sup>a</sup> (n=527) |  | No (n=381) |  | Total (N=908) |  |  |
|  |  | n | % | n | % | n | % |  |
| Study region | Guadalajara | 182 | 35 | 120 | 32 | 302 | 33 | <0.001 |
|  | La Rioja | 135 | 26 | 61 | 16 | 196 | 22 |  |
|  | Basque Country | 210 | 40 | 200 | 52 | 410 | 45 |  |
| Marital status | Married /Civil Partnership | 83 | 16 | 164 | 43 | 247 | 27 | <0.001 |
|  | Single | 441 | 84 | 203 | 53 | 644 | 71 |  |
|  | Divorced | 3 | 0.6 | 14 | 3.7 | 17 | 1.9 |  |
| Origin | Spain | 460 | 87 | 167 | 44 | 627 | 69 | <0.001 |
|  | Latin America | 50 | 10 | 176 | 46 | 226 | 25 |  |
|  | Rest of the world<sup>c</sup> | 17 | 3.2 | 38 | 10 | 55 | 6.1 |  |
| Previous screening<sup>d</sup> | Yes | 426 | 81 | 296 | 78 | 722 | 80 | 0.25 |
| Sexual orientation | Heterosexual | 475 | 90 | 351 | 92 | 826 | 91 | 0.29 |
|  | Bisexual | 38 | 7.2 | 18 | 4.7 | 56 | 6.2 |  |
|  | Homosexual | 14 | 2.7 | 12 | 3.1 | 26 | 2.9 |  |
| Sexual partners | 1–5 | 311 | 59 | 252 | 66 | 563 | 62 | 0.011 |
|  | 6–10 | 135 | 26 | 95 | 25 | 230 | 25 |  |
|  | >10 | 81 | 15 | 34 | 8.9 | 115 | 13 |  |
| Condom use | Always | 149 | 28 | 69 | 18 | 218 | 24 | <0.001 |
|  | Most of the time | 100 | 19 | 52 | 14 | 152 | 17 |  |
|  | Half of the time | 25 | 4.7 | 24 | 6.3 | 49 | 5.4 |  |
|  | Occasionally | 75 | 14 | 71 | 19 | 146 | 16 |  |
|  | Never | 178 | 34 | 165 | 43 | 343 | 38 |  |
| Sexually transmitted Infection (STI) | Any STI | 37 | 7.0 | 23 | 6.0 | 60 | 6.6 | 0.56 |
|  | Chlamydia infection | 13 | 2.5 | 9 | 2.4 | 22 | 2.4 | 0.92 |
|  | Genital herpes | 14 | 2.7 | 7 | 1.8 | 21 | 2.3 | 0.42 |
|  | Genital warts | 7 | 1.3 | 7 | 1.8 | 14 | 1.5 | 0.54 |
|  | Trichomoniasis | 3 | 0.57 | 4 | 1.1 | 7 | 0.77 | 0.33 |
|  | Syphilis | 1 | 0.19 | 0 |  | 1 | 0.11 | 0.58 |
|  | Gonorrhea | 1 | 0.19 | 0 |  | 1 | 0.11 | 0.58 |
|  | HIV infection | 0 |  | 0 |  | 0 |  |  |
| Parity | 0 | 456 | 87 | 222 | 58 | 678 | 75 | <0.001 |
|  | 1 | 50 | 10 | 89 | 23 | 139 | 15 |  |
|  | ≥2 | 21 | 4.0 | 70 | 18 | 91 | 10 |  |
| Smoking status | Smoker | 109 | 21 | 68 | 18 | 177 | 19 | 0.29 |
|  |  | Mean | SD<sup>e</sup><br>± | Mean | SD ± | Mean | SD ± |  |
| Age (years) |  | 28.9 | 1.5 | 30.1 | 1.7 | 29.4 | 1.7 | <0.001 |
| Age sexual debut |  | 17.4 | 2.4 | 17.7 | 2.9 | 17.6 | 2.6 | 0.35 |
| Years of smoking |  | 10.8 | 3.9 | 11.6 | 5.4 | 11.1 | 4.5 | 0.11 |

A: Women who received ≥1 dose of any HPV vaccine.
b: Statistically significant results are shown in bold.
c: Includes 26 women from Europe, 28 from Africa and 6 from other regions.
d: Tested with any screening test in her lifetime.
e: SD: Standard Deviation.

Most of the women were vaccinated within routine regional immunization schedule (88%; 463/527), mainly with the 4vHPV (87%; 460/527). However, some participants were vaccinated outside the Spanish regions included in the study, either abroad (3.4%; 18/527) or in other Spanish regions (5.7%; 30/527). In addition, 12% (61/527) of women were vaccinated after sexual initiation. No information on the type of vaccine was available in 7.2% (38/527) (Table 2).

**Table 2:**
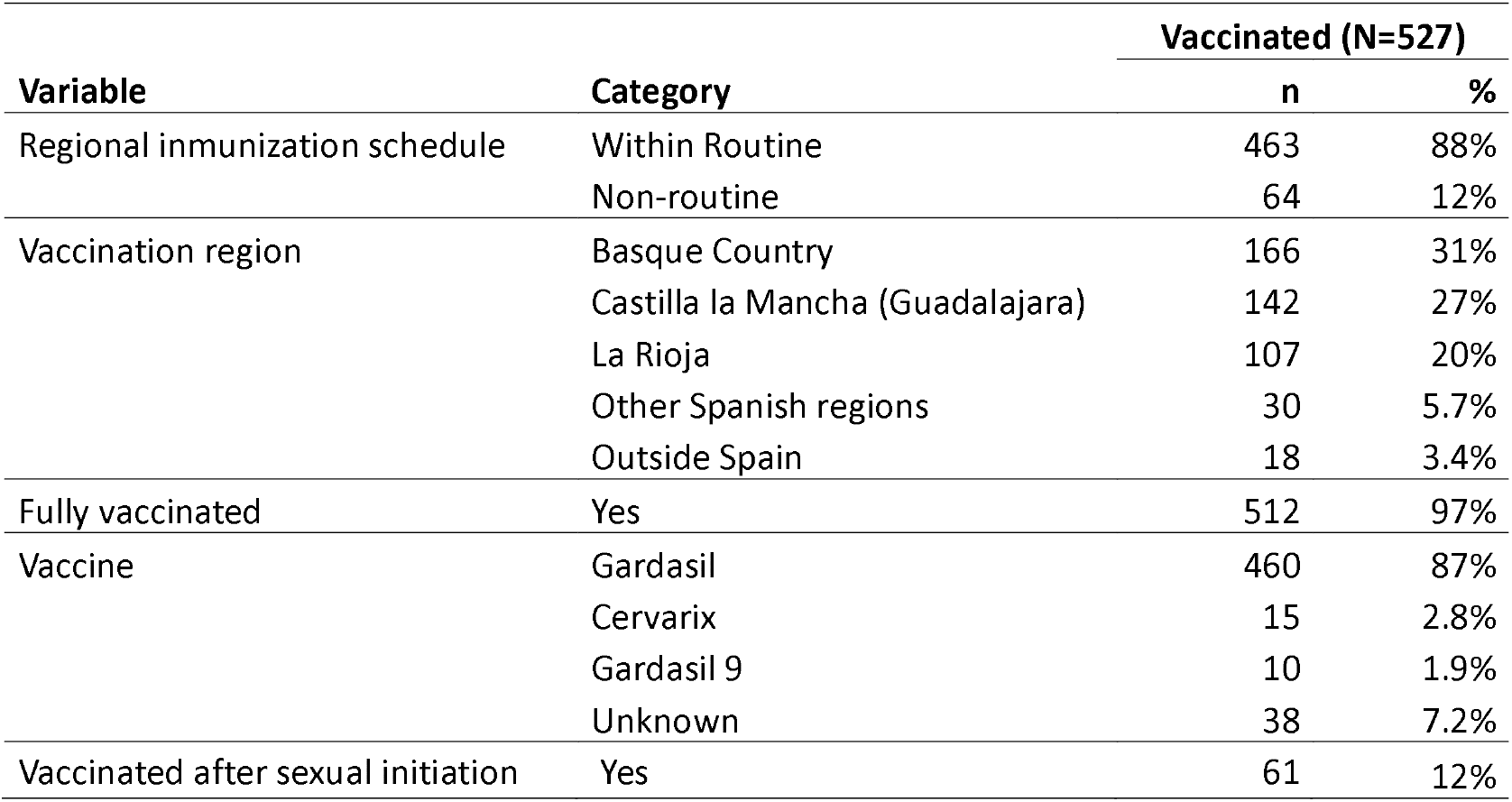
Participants vaccination data.

| Variable | Category | Vaccinated (N=527) |  |
| --- | --- | --- | --- |
|  |  | n | % |
| Regional immunization schedule | Within Routine | 463 | 88% |
|  | Non-routine | 64 | 12% |
| Vaccination region | Basque Country | 166 | 31% |
|  | Castilla la Mancha (Guadalajara) | 142 | 27% |
|  | La Rioja | 107 | 20% |
|  | Other Spanish regions | 30 | 5.7% |
|  | Outside Spain | 18 | 3.4% |
| Fully vaccinated | Yes | 512 | 97% |
| Vaccine | Gardasil | 460 | 87% |
|  | Cervarix | 15 | 2.8% |
|  | Gardasil 9 | 10 | 1.9% |
|  | Unknown | 38 | 7.2% |
| Vaccinated after sexual initiation | Yes | 61 | 12% |

### HPV genotype distribution differs according to vaccination status

All cervical samples tested for HPV yielded valid results, with a mean of 532 (SD ± 15) thousand human cells per PCR reaction. HPV was detected in 37% (337/908) of the samples. Co-detection of multiple HPV genotypes was observed in 18% (163/908) of cases, and HR-HPV was present in 24% (221/908). No statistically significant differences were found between VW and UVW (Table 3).

**Table 3:** HPV and cytology test results according vaccination status.

| Test | Category<sup>a</sup> | Vaccination |  |  |  |  |  | *p*-value<sup>b</sup> |
| --- | --- | --- | --- | --- | --- | --- | --- | --- |
|  |  | Yes (n=527) |  | No (n=381) |  | Total (N=908) |  |  |
|  |  | n | % | n | % | n | % |  |
| HPV | HPV positive | 194 | 37 | 143 | 38 | 337 | 37 | 0.82 |
|  | Single detection | 101 | 19 | 73 | 19 | 174 | 19 | 0.99 |
|  | Co-detection | 93 | 18 | 70 | 18 | 163 | 18 | 0.78 |
|  | HR-HPV | 120 | 23 | 101 | 27 | 221 | 24 | 0.19 |
| Cytology <sup>c</sup> | NILM | 484 | 92 | 346 | 91 | 830 | 92 | 0.52 |
|  | ASCUS | 19 | 3.6 | 16 | 4.2 | 35 | 3.9 | 0.65 |
|  | LSIL | 21 | 4.0 | 10 | 2.6 | 31 | 3.4 | 0.26 |
|  | ASC-H | 1 | 0.19 | 2 | 0.52 | 3 | 0.33 | 0.58 |
|  | HSIL | 1 | 0.19 | 6 | 1.6 | 7 | 0.77 | **0.046** |
|  | AGC-FN | 0 | 0 | 1 | 0.26 | 1 | 0.11 | 0.42 |
|  | ASC-H/AGC-FN/HSIL | 2 | 0.38 | 9 | 2.4 | 11 | 1.2 | **0.011** |

a: AGC-FN: Atypical glandular cells -favor neoplastic; ASCUS: Atypical squamous cells of undetermined significance; LSIL: Low-grade squamous intraepithelial lesion; ASC-H: Atypical squamous cells, cannot exclude HSIL; HSIL: High-grade squamous intraepithelial lesion; NILM: Negative for intraepithelial lesion or malignancy. HR-HPV: High-risk HPV.
b: Statistically significant results are shown in bold.
c: One cytology test from a vaccinated women was invalid and was not included in the statistical analysis.

Regarding genotype distribution, HPV 16 was more frequent in UVW than in VW (4.2% vs 0.76%; p=0.001). Similarly, HPV 31 (3.4% vs 1.3%; p=0.035) and HPV 18 (1.3% vs 0%; p=0.013), the latter being detected only in UVW, also showed a higher frequency among UVW (Table 4). In contrast, HPV 51 (3.2% vs 6.3%; p=0.033) and HPV 70 (0.78% vs 2.7%; p=0.032) were less frequently detected in UVW than in VW (Table 4). Other genotypes included in the 4vHPV did not show statistically significant differences between UVW and VW: HPV 6 (0.79% vs 0.38%; p=0.352) and HPV 11 (0.52% vs 0%; p=0.176), although HPV 11 was only detected in UVW (Table 4). VL of the different genotypes did not show statistical differences between both VW and UVW, except for HPV 16 (see below and Supplementary material).

**Table 4:**
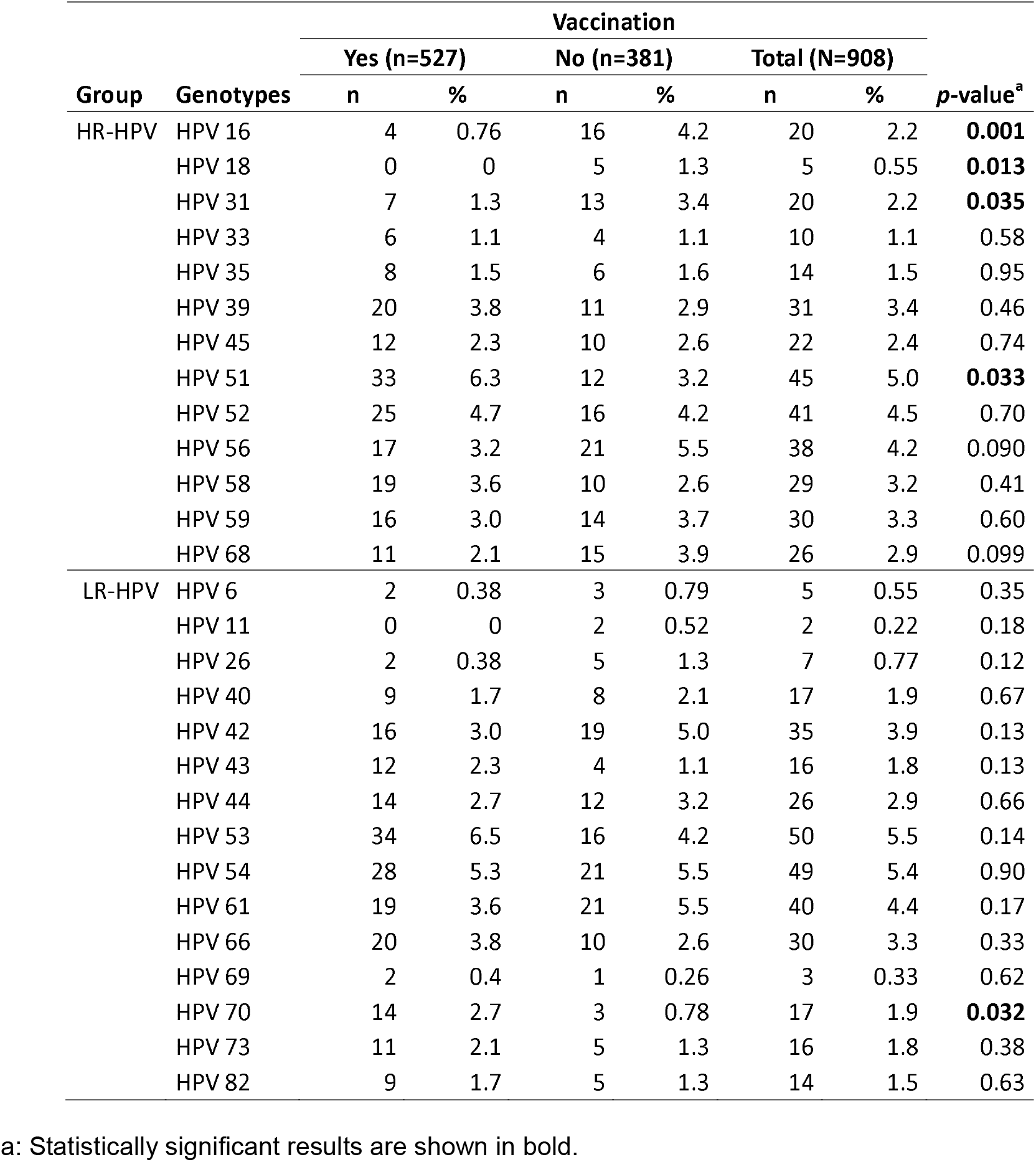
Distribution of HPV genotypes according to the vaccination status.

### Vaccine-targeted HPV genotypes and genital warts in vaccinated women

Four cases of HPV 16 were detected in fully VW with 4vHPV, two of whom had been vaccinated after the initiation of sexual activity. Notably, these participants showed a low VL, particularly those vaccinated before sexual debut (0.08 and 0.19 virus/1000 human cells) compared to those after sexual initiation (0.43 and 299 virus/1000 human cells). Moreover, the VL of HPV 16 in VW was lower than in UVW (mean log VL: 0.08 ± SD 1.6 vs 2.4 ± SD 1.7 copies/1000 human cells; p=0.050) (Supplementary table 1).

The two HPV 6 cases detected among VW corresponded to one participant who had received the 9vHPV after sexual debut, and to a Latin American woman fully vaccinated in her country with an unknown vaccine; thus, it cannot be ruled out that she could be vaccinated with 2vHPV.

History of genital warts was reported in 1.3% (7/527) of VW (Table 1). Among these cases, five women had been fully vaccinated after sexual debut (four with 4vHPV vaccine and one with an unknown vaccine), one had received 2vHPV before sexual debut, and only one had been vaccinated with 4vHPV prior to sexual initiation, thus having a potential optimal protection against genital warts.

Four of the seven VW with detected HPV 31 had received the vaccine after sexual debut (two with 4vHPV and two with 9vHPV). The remaining three women had been vaccinated before sexual debut (two with 4vHPV and one with an unknown vaccine type) and could therefore have benefited from vaccine cross-protection. The three of them showed low VL (0.001, 2, and 7 copies per 1,000 human cells).

### Decrease of cytological abnormalities in vaccinated women

Valid cytological test results were available for 907 samples. Abnormal cytological results were observed in 8.5% (77/907) of the participants: 3.9% (35/907) atypical squamous cells of undetermined significance (ASCUS), 3.4% (31/907) low grade squamous intraepithelial lesion (LSIL), and 1.2% (11/907) high-grade lesions. The latter includes: high grade squamous intraepithelial lesion (HSIL), atypical squamous cells which cannot exclude HSIL (ASC-H), and atypical glandular cells-favor neoplastic (AGC-FN) (Table 3). The frequency of high-grade lesions (ASC-H/AGC-FN/HSIL) was higher in the UVW than in VW (2.4% vs 0.38%, p = 0.011). A similar pattern was observed for HSIL when analyzed individually (1.6% vs 0.19%, p = 0.046) (Table 3).

### Vaccination status, epidemiological and sexual behavior factors are associated with the detection of HPV genotypes

A multivariable logistic regression analysis was performed to identify variables independently associated with their detection. The presence of HPV 16/18 was significantly associated with vaccination status, with UVW being nearly five times more likely to detect HPV 16/18 compared with VW [adjusted odds ratio (aOR): 4.9; 95% CI: 1.4–17]. This association was even stronger considering only 4vHPV (aOR: 8.7; 95% CI: 1.8–43). Vaccination after the sexual debut was also identified as associated factor (aOR: 8.0; 95% CI: 1.6-39). HPV 16/18 detection was also related to cytological abnormalities, particularly with more severe lesions. Women with high-grade lesions (ASC-H/AGC-FN/HSIL), compared to those negative for intraepithelial lesions or malignancy (NILM), were 13 times more likely to have HPV 16/18 detected (aOR: 13; 95% CI: 2.9–58) (Figure 1 and Supplementary Table 2).

**Figure 1.**
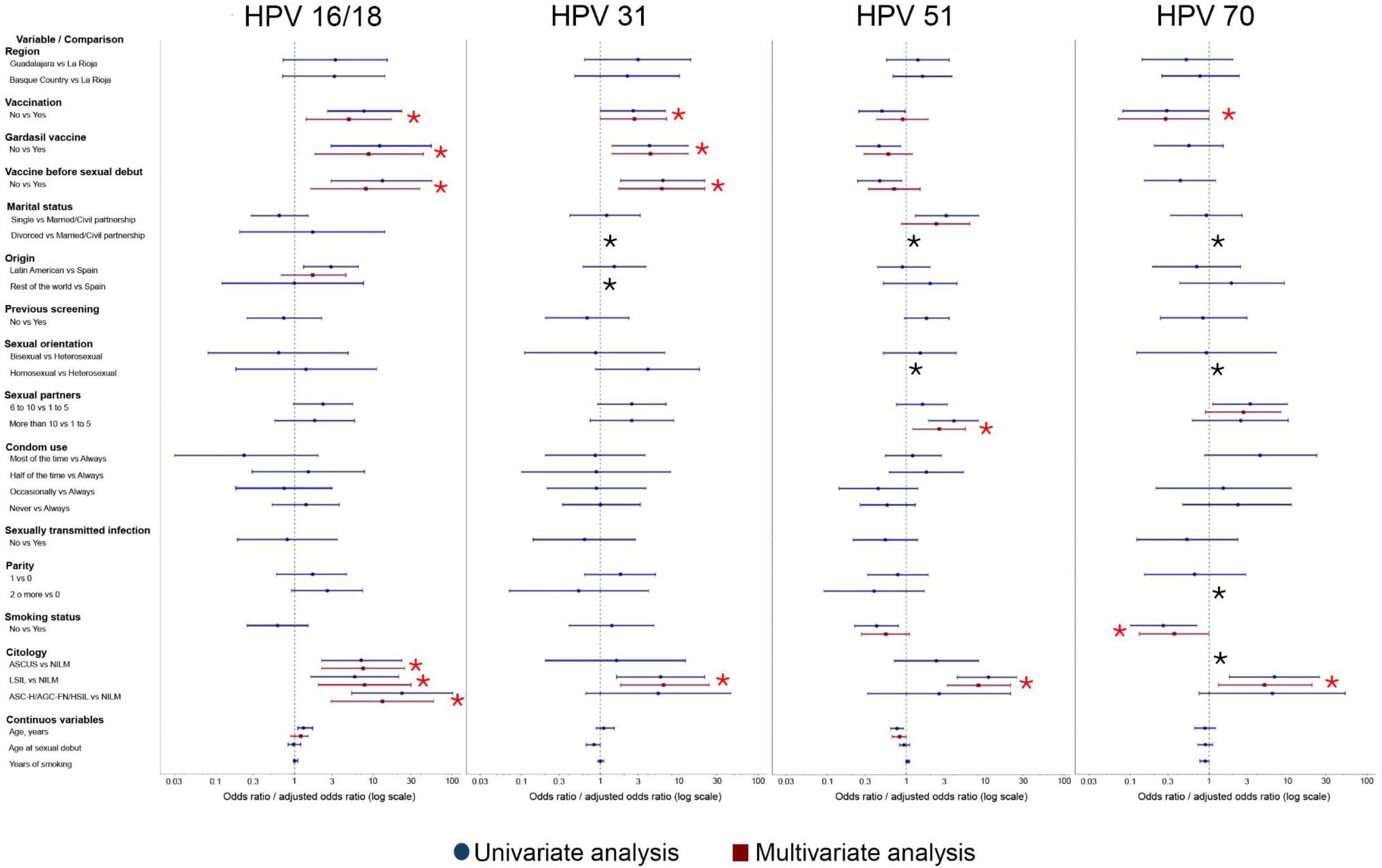
Forest plots showing factors associated with the detection of different HPV genotypes. Points and squares represent odds ratios, and horizontal lines represent 95% confidence intervals, plotted on a logarithmic scale. Odds ratios that could not be estimated because of zero observations in one of the comparison categories are indicated by black asterisks. Statistically significant associations in the multivariable analysis are indicated by red asterisks.

HPV 31 detection was also associated with vaccination status and cytological abnormalities. HPV 31 was more likely to be detected in UVW (aOR: 2.7; 95% CI: 1.0– 6.9), with a stronger effect observed among those vaccinated with 4vHPV (aOR: 4.3; 95% CI: 1.4–13) and also associated with vaccination after sexual initiation (aOR: 6.0; 95% CI: 1.7-21) and with the presence of LSIL (aOR: 6.3; 95% CI: 1.8–24) (Figure 1 and Supplementary table 3).

In contrast, HPV 51 detection was not associated with vaccination status (aOR: 0.90; 95% CI: 0.42–1.9). HPV51 was, however, associated with a higher number of lifetime sexual partners (>10 vs 1 to 5) (aOR: 2.6; 95% CI: 1.2–5.6), and having LSIL compared to NILM (aOR: 8.2; 95% CI: 3.3–21).

HPV 70 showed a borderline association with vaccination status (aOR: 0.28; 95% CI: 0.07–1.0), being more likely to be detected among VW, although it was not observed if considering only 4vHPV-vaccinated women. However, HPV 70 detection was associated with LSIL (aOR: 5.0; 95% CI: 1.3–20), while non-smoking appeared to have a protective effect (aOR: 0.36; 95% CI: 0.13–0.99) (Figure 1 and Supplementary Table 4 and 5).

## DISCUSSION

This study characterizes a cohort of women participating in population-based cervical cancer screening programs in three Spanish regions and evaluates the impact of HPV vaccination on HPV genotype distribution and cytological abnormalities.

Our results demonstrate a substantial reduction in vaccine-targeted HPV genotypes among VW. HPV 16/18 were significantly more frequent in UVW, and the odds of detecting them were almost nine times higher when comparing 4vHPV-vaccinated women. These findings are particularly relevant because HPV 16/18 account for approximately 77% of cervical cancers worldwide^6^. The observed reductions are consistent with evidence from clinical trials and population-based studies showing major declines in vaccine-type HPV infections following the introduction of vaccination^18-24^. Similar findings have also been reported in previous studies conducted in Spain^14,15^.

In our study, HPV 16/18 detection was strongly associated with high-grade cytological abnormalities, supporting the clinical relevance of the reduction observed among VW. In agreement with this finding, high-grade lesions were significantly less frequent in VW than in UVW, providing early evidence of vaccine effectiveness in preventing precancerous cervical disease.

HPV 16 was detected in a small number of VW. Some of these cases could be explained by vaccination after sexual debut, a factor independently associated with HPV 16/18 detection in our study and known to reduce vaccine effectiveness^25^. Interestingly, women vaccinated before sexual initiation showed very low HPV16 VL. These detections may represent transient viral deposition rather than true persistent infection. Further studies for clarifying the nature and clinical relevance of these detections could be particularly important for avoiding unnecessary referrals and interventions when vaccine-targeted genotypes are detected in vaccinated women before the sexual debut.

Cross-protection against phylogenetically related HPV genotypes has been described for both 2vHPV and 4vHPV vaccines, particularly for HPV 31^24,26,27^ and associated with antibodies against HPV 16^28^. In our study, HPV 31 was significantly less frequent among VW, supporting the existence of cross-protective effects in routine clinical practice. Similar to HPV 16, several HPV 31 detections in VW before sexual initiation were characterized by low VL, which could indicate transient viral deposition.

The frequencies of HPV 6, HPV 11 and genital warts were low among VW although not statistically significant. Most VW reporting genital warts had been vaccinated after sexual debut or had received a vaccine not covering HPV 6/11. Only one VW with genital warts had completed the full 4vHPV schedule before sexual debut. However, about 10% of genital warts are caused by non-HPV 6/11 genotypes^29^. Overall, these findings remain consistent with the established protective effect of HPV vaccination against HPV 6/11-related disease^30-32^, including some studies from Spain^11-14^.

HPV 51 and HPV 70 were more frequently detected among VW in the unadjusted analysis. However, multivariable models did not support an association with vaccination status. Instead, HPV 51 was associated with a higher number of sexual partners and LSIL, whereas HPV 70 was associated with smoking status and LSIL. Both number of sexual partners and smoking are well known risk factors for HPV infections^33^. These findings suggest that differences in exposure rather than vaccination explain the observed genotype distribution.

Potential concerns following vaccine implementation include genotype replacement and clinical unmasking^34^. However, our results do not support either phenomenon, as vaccination was not independently associated with increased detection of non-vaccine HPV genotypes. Nevertheless, further longitudinal studies investigating the etiology of cervical lesions and integrating epidemiological, behavioral and clinical factors are needed to determine whether these potential phenomena do actually occur in the population and their clinical relevance.

Compared with an age-matched pre-vaccination cohort in Spain (CLEOPATRE study 2008)^35^, VW showed a markedly lower prevalence of HPV 16/18 (0.76% vs. 4.3%) consistent with the protective effect of vaccination. However, we observed a higher HPV detection rate between the post and the pre-vaccination era (37% vs. 19%). These comparisons should be interpreted cautiously because our cohort reported earlier sexual debut (17.6 vs. 19.8 years) and a higher number of lifetime sexual partners (>5 partners: 38% vs. 10%) than historical data from the same age cohort (AFRODITA study 2005)^36^, indicating greater level of sexual activity in the current Spanish population and increased cumulative exposure to HPV. Similarly, HPV 51, one of the most frequently detected HPV genotypes in several countries^14,37-41^ and in the present study, increased from 1.3% in the pre-vaccination era (CLEOPATRE study)^35^ to 6.3% among VW in our cohort, probably reflecting increased sexual activity rather than vaccine-related genotype replacement. Despite its low oncogenic contribution^6,7^, continued surveillance is warranted to monitor its potential clinical impact in the post-vaccination era.

Our study illustrates the complexity of evaluating vaccine effectiveness in the real-world setting. The study population included VW with different vaccine types, individuals vaccinated outside the regional immunization programs, and participants with incomplete vaccination records. In addition, approximately 12% of VW received the vaccine after sexual debut, an important determinant of vaccine effectiveness^25^. Besides, VW and UVW groups have differences that can distort the observed vaccine-impact if they are not considered, as illustrated by the apparently higher prevalence of HPV 51 and HPV 70 among VW in our cohort.

The lower prevalence of HR-HPV genotypes and severe cytological abnormalities observed among VW in this study have important implications for cervical cancer screening. These findings support risk-based screening strategies, including primary HPV testing and vaccination-tailored age at screening initiation^42-44^, recently adopted in Spain^45,46^ with the Basque Country being the first Spanish region to implement it in 2026. Such strategies may improve screening efficiency as vaccinated cohorts enter screening age. Moreover, the higher proportion of migrant women among the UVW further underscores the need to strengthen targeted vaccination and screening strategies for this vulnerable population.

Several limitations should be considered. The study was conducted in three regions where vaccination was predominantly based on 4vHPV, limiting comparisons according to vaccine type. Larger studies including regions with different vaccination strategies will be required to evaluate less prevalent genotypes and assess the impact of 9vHPV, the newest HPV vaccine introduced. The vaccination status of some participants with relied partly on self-reported information, particularly among mobile participants and due to the limited interoperability between regional vaccination and screening databases.

In conclusion, this multicenter study combines comprehensive HPV genotyping, viral load quantification, cytological assessment, and detailed vaccination, epidemiological, and sexual behavior data, enabling a robust evaluation of vaccine effectiveness while accounting for key confounding factors. This integrated approach provides a detailed characterization of genotype-specific HPV distribution in the post-vaccination era, allowing the assessment of direct vaccine effects, cross-protection, and the potential influence of epidemiological and behavioral risk factors. These findings support the need for continuous HPV molecular surveillance to monitor vaccine impact, optimize screening strategies, and improve understanding of HPV epidemiology in vaccinated populations.

## Data Availability

All data produced in the present study are available upon reasonable request to the authors

## AUTHOR CONTRIBUTIONS

Conceptualization: E.A.L, J.W.C.M., D.V.P, R.R.T, M.J.P.M., H.G.; Data curation: H.G.; Formal analysis: A.I.G.S., H.G.; Funding acquisition: H.G.; Investigation: A.I.G.S., A.D.E., M.A.M., B.D.E., M.C.A., T.Z.F., C.R.S., Spanish HPV Vaccine Impact Study Group; Methodology: A.I.G.S.; Project administration: D.V.P., R.R.T., M.J.P.M.; Resources: M.A.M., S.P.C., B.D.E., M.C.A., T.Z.F., J.S.G., Spanish HPV Vaccine Impact Study Group; Supervision: E.A.L., A.D.E., S.P.C., B.D.E., M.C.A., J.S.G.; Validation: M.A.M., J.W.C.M., S.P.C., T.Z.F., J.S.G., H.G.; Writing-original draft: A.I.G.S.; Writing-review and Editing: E.A.L, J.W.C.M., D.V.P., R.R.T., M.J.P.M., C.R.S., H.G.

## ACKNOWLEDGMENTS

We thank all participants in the cervical cancer screening programs in La Rioja, Guadalajara, and the Basque Country who agreed to participate in this study. We also thank Xavier Bosch and Esther Roura from the Institut Català d’Oncologia ICO for sharing data from the CLEOPATRE study, and Miguel Thomson and Elena Delgado from the HIV Biology and Variability Unit at CNM-ISCIII for their support. We are also grateful to Asunción Díaz Franco from the HIV, STI and Hepatitis Surveillance Unit (National Epidemiology Center, ISCIII) for her advice on the statistical analysis and the Information and Communications Technology Unit (UTIC) from the ISCIII for the design and management of the online questionnaire.

## MEMBERS OF THE SPANISH HPV VACCINE IMPACT STUDY GROUP

### Guadalajara Health Area

Raquel Alonso Cabanillas, Raquel Castillo Castillo, Alicia Blanco García, Rosa María Fernández Turiño, Rocío García Pagola, María Dolores Garrido Moreno, Sarai Gordo Hidalgo, Javier Jiménez Carrera, Asunción Las Heras López, María Jesús de Luis López, Olga Marta Nova Gorro, María Cortes Sancha Sánchez, Noelia Sanz del Pozo, Gima Varona Esquivel. **Hospital Universitario San Pedro (La Rioja):** Mariano Laguna Olmo and Belén Merino García. **Basque Health Service:** Rosa Alonso Tofe, María Asunción Arrieta Ugarte, Lía Belinda Ataupillco Remuzgo, Idoia Briones Pérez, Virginia Campillo Azkuenaga, Uxue Castellano Etxeberria, Rosa María Castro García, Cristina Castro Rodríguez, Natalia Cobo García, Pilar Costillas Caño, Irene del Campo Fuentes, Ziortza Echebarria Vadillo, Laura Falcón Carvajal, Janire García Reina, Garazi González Corrales, Ane Guilló Arakistain, Oihane Herrero Ibáñez, Ane Izaguirre Díaz, Iris Izurza Elizondo, Susana Jorrín Cuesta, Rocío Lagares García, Amaia López de Maturana Elorza, Idoya López Muñiz, Irati Marín Salazar, Patricia Martínez López, June Martínez Muñoz, Tania Mateo Urdiales, Lucía Monzón Muñoz, María Nieves Gonzalez Cordero, Ana Ortega Sánchez, Ane Pascual Díaz, David Redondo Collado, Francisco Javier Rey Bravo, Laura Rodríguez Riesco, María Begoña Roncero Santos, María Sala Higón, Eva Santaolalla Pascual, Arabela Vidal Ivars, Ane Vitales Retegi and Lara Delia Zuazo González. **CNM-ISCIII (Madrid):** Manuela Rodríguez Vargas. **Hospital Central de la Defensa Gómez Ulla “CSVE” (Madrid):** María Simón Sacristán.

## DATA AVAILABILITY

Anonymized data may be available from the corresponding author upon reasonable formal request.

## SUPPLEMENTARY INFORMATION

**Supplementary material table 1:** Genotype viral load according vaccination status.

**Supplementary material table 2:** Factors associated with the detection of HPV 16/18.

**Supplementary material table 3:** Factors associated with the detection of HPV 31.

**Supplementary material table 4:** Factors associated with the detection of HPV 51.

**Supplementary material table 5:** Factors associated with the detection of HPV 70.

**Questionnaire**

