## Supplementary material for "Impact of HPV vaccination in Spain: Reductions in vaccine-type genotypes, cytological abnormalities, and the influence of epidemiological factors": suplemntary material

|  | **Vaccination** | | | | | |  |
| --- | --- | --- | --- | --- | --- | --- | --- |
|  | **Yes** | | | **No** | | |  |
| **Genotype** | **n** | **Mean**  **log VL** | **SD±^a^** | **n** | **Mean**  **log VL** | **SD±^a^** | ***p*-value^c^** |
| HPV 6 | 2 | 0.18 | 0.54 | 3 | 0.63 | 2.5 | 0.80 |
| HPV 11 | 0 | NA^b^ | NA | 2 | 1.6 | 1.8 | NA |
| HPV 16 | 4 | 0.08 | 1.6 | 16 | 2.4 | 1.7 | **0.050** |
| HPV 18 | 0 | NA | NA | 5 | -2.3 | 1.0 | NA |
| HPV 31 | 7 | 1.0 | 2.4 | 13 | 0.8 | 2.2 | 0.89 |
| HPV 33 | 6 | 2.5 | 2.1 | 4 | 3.5 | 1.3 | 0.61 |
| HPV 35 | 8 | 2.1 | 2.0 | 6 | 0.79 | 1.4 | 0.23 |
| HPV 39 | 20 | -0.62 | 2.0 | 11 | -0.02 | 2.5 | 0.54 |
| HPV 45 | 12 | 0.71 | 2.0 | 10 | 2.1 | 1.4 | 0.15 |
| HPV 51 | 33 | 1.2 | 2.3 | 12 | 0.87 | 2.0 | 0.67 |
| HPV 52 | 25 | 1.3 | 1.9 | 16 | 1.8 | 2.3 | 0.38 |
| HPV 56 | 17 | 0.59 | 2.1 | 21 | -0.21 | 1.9 | 0.34 |
| HPV 58 | 19 | 0.97 | 2.0 | 10 | 0.72 | 2.0 | 0.88 |
| HPV 59 | 16 | 0.23 | 1.9 | 14 | 0.48 | 1.9 | 0.55 |
| HPV 68 | 11 | 0.73 | 2.3 | 15 | -0.12 | 1.4 | 0.20 |

**Supplementary material table 1:** Genotype viral load according vaccination status

Viral load is quantified as the number of copies per 1,000 human cells and reported on a logarithmic scale

a: SD: Standard deviation.

b: NA: Not applicable.

c. Statistically significant results are shown in bold.

|  |  | **HPV 16/18** | | | | **Univariant ^a^** | | **Multivariant ^a^** | |
| --- | --- | --- | --- | --- | --- | --- | --- | --- | --- |
|  |  | **No detected** | | **Detected** | |  |  |  |  |
| **Variable** | **Category** | **n** | **%** | **n** | **%** | **OR** | **95% CI** | **aOR** | **95% CI** |
| Region | La Rioja | 194 | 22 | 2 | 8 | Reference | | Reference | |
|  | Guadalajara | 292 | 33 | 10 | 40 | 3.3 | 0.72-15 |  |  |
|  | Basque Country | 397 | 45 | 13 | 52 | 3.2 | 0.71-14 |  |  |
| Vaccination | Yes | 523 | 59 | 4 | 16 | Reference | | Reference | |
|  | No | 360 | 41 | 21 | 84 | **7.6** | **2.6-23** | **4.9** | **1.4-17** |
| Gardasil vaccine | Yes | 458 | 52 | 2 | 8 | Reference | | Reference | |
|  | No | 425 | 48 | 23 | 92 | **12** | **2.9-54** | **8.7** | **1.8-43** |
| Vaccine before | Yes | 464 | 53 | 23 | 92 | Reference | | Reference | |
| Sexual debut | No | 419 | 47 | 2 | 8 | **13** | **2-9-55** | **8.0** | **1.6-39** |
| Marital status | Married/Civil Partnership | 238 | 27 | 9 | 36 | Reference | | Reference | |
|  | Single | 629 | 71 | 15 | 60 | 0.64 | 0.28-1.5 |  |  |
|  | Divorced | 16 | 1.8 | 1 | 4 | 1.7 | 0.20-14 |  |  |
| Origen | Spain | 615 | 70 | 12 | 48 | Reference | | Reference | |
|  | Latin America | 214 | 24 | 12 | 48 | **2.9** | **1.3-6.5** | 1.7 | 0.68-4.5 |
|  | Rest of the world | 54 | 6.1 | 1 | 4 | 1.0 | 0.12-7.5 |  |  |
| Previous screening | Yes | 701 | 79 | 21 | 84 | Reference | | Reference | |
|  | No | 182 | 21 | 4 | 16 | 0.73 | 0.25-2.2 |  |  |
| Sexual orientation | Heterosexual | 803 | 91 | 23 | 92 | Reference | | Reference | |
|  | Bisexual | 55 | 2.8 | 1 | 4 | 0.63 | 0.08-4.8 |  |  |
|  | Homosexual | 25 | 6.2 | 1 | 4 | 1.4 | 0.18-11 |  |  |
| Sexual partners | 1 to 5 | 552 | 63 | 11 | 44 | Reference | | Reference | |
|  | 6 to 10 | 220 | 25 | 10 | 40 | 2.3 | 0.96-5.5 |  |  |
|  | >10 | 111 | 13 | 4 | 16 | 1.8 | 0.56-5.8 |  |  |
| Condom Use | Always | 212 | 24 | 6 | 24 | Reference | | Reference | |
|  | Most of the time | 151 | 17 | 1 | 4 | 0.23 | 0.03-2.0 |  |  |
|  | Half of the time | 47 | 5.3 | 2 | 8 | 1.5 | 0.29-7.7 |  |  |
|  | Occasionally | 143 | 16 | 3 | 12 | 0.74 | 0.18-3.0 |  |  |
|  | Never | 330 | 37 | 13 | 52 | 1.4 | 0.52-3.7 |  |  |
| Sexual Transmitted | Yes | 58 | 6.6 | 2 | 8 | Reference | | Reference | |
| Infection | No | 825 | 93 | 23 | 92 | 0.81 | 0.19-3.5 |  |  |
| Parity | 0 | 663 | 75 | 15 | 60 | Reference | | Reference | |
|  | 1 | 134 | 15 | 5 | 20 | 1.7 | 0.59-4.6 |  |  |
|  | ≥ 2 | 86 | 10 | 5 | 20 | 2.6 | 0.91-7.3 |  |  |
| Smoking status | Yes | 170 | 19 | 7 | 28 | Reference | | Reference | |
|  | No | 713 | 81 | 18 | 72 | 0.61 | 0.25-1.5 |  |  |
| Cytology | NILM | 815 | 93 | 15 | 60 | Reference | | Reference | |
|  | ASCUS | 31 | 3.5 | 4 | 16 | **7** | **2.2-23** | **7.4** | **2.2-25** |
|  | LSIL | 28 | 3.2 | 3 | 12 | **5.8** | **1.6-21** | **7.7** | **2.0-30** |
|  | ASC-H/AGC_FN/HSIL | 7 | 0.8 | 3 | 12 | **23** | **5.3-102** | **13** | **2.9-58** |
| **Variable** |  | **n** | **Mean** | **n** | **Mean** |  |  |  |  |
| Age (years) |  | 883 | 29,4 | 25 | 30,2 | **1.3** | **1.1-1.7** | 1.2 | 0.89-1.5 |
| Age sexual debut |  | 883 | 17,6 | 25 | 17,4 | 0.97 | 0.83-1.2 |  |  |
| Years of smoking |  | 166 | 11 | 6 | 13,5 | 1.0 | 0.98-1.1 |  |  |

**Supplementary material table 2:** Factors associated with the detection of HPV 16/18

a: statistically significant results are shown in bold. p value provided

|  |  | **HPV 31** | | | | **Univariant ^a^** | | **Multivariant ^a^** | |
| --- | --- | --- | --- | --- | --- | --- | --- | --- | --- |
|  |  | **No detected** | | **Detected** | |  |  |  |  |
| **Variable** | **Category** | **n** | **%** | **n** | **%** | **OR** | **95% CI** | **aOR** | **95% CI** |
| Region | La Rioja | 194 | 22 | 2 | 10 | Reference | | Reference | |
|  | Guadalajara | 293 | 33 | 9 | 45 | 3.0 | 0.63-14 |  |  |
|  | Basque Country | 401 | 45 | 9 | 45 | 2.2 | 0.47-10 |  |  |
| Vaccination | Yes | 520 | 59 | 7 | 35 | Reference | | Reference | |
|  | No | 368 | 41 | 13 | 65 | **2.6** | **1.0-6.7** | **2.7** | **1.0-6.9** |
| Gardasil4 vaccine | Yes | 456 | 51 | 4 | 20 | Reference | | Reference | |
|  | No | 432 | 49 | 16 | 80 | **4.2** | **1.4-13** | **4.3** | **1.4-13** |
| Vaccination before | Yes | 463 | 52 | 3 | 15 | Reference | | Reference | |
| sexual debut | No | 425 | 48 | 17 | 85 | **6.2** | **1.8-21** | **6.0** | **1.7-21** |
| Marital status | Married/Civil Partnership | 242 | 27 | 5 | 25 | Reference | | Reference | |
|  | Single | 629 | 71 | 15 | 75 | 1.2 | 0.41-3.2 |  |  |
|  | Divorced | 17 | 2 | 0 | 0 | NA | *p=* 0.52 |  |  |
| Origen | Spain | 614 | 69 | 13 | 65 | Reference | | Reference | |
|  | Latin America | 219 | 25 | 7 | 35 | 1.5 | 0.60-3.8 |  |  |
|  | Rest of the world | 55 | 6.2 | 0 | 0 | NA | *p=* 0.28 |  |  |
| Previous screening | Yes | 705 | 79 | 17 | 85 | Reference | | Reference | |
|  | No | 183 | 21 | 3 | 15 | 0.68 | 0.20-2.3 |  |  |
| Sexual orientation | Heterosexual | 809 | 91 | 17 | 85 | Reference | | Reference | |
|  | Bisexual | 55 | 6.2 | 1 | 5 | 0.87 | 0.11-6.6 |  |  |
|  | Homosexual | 24 | 3 | 2 | 10 | 4.0 | 0.86-18 |  |  |
| Sexual partners | 1-5 | 555 | 63 | 8 | 40 | Reference | | Reference | |
|  | 6-10 | 222 | 25 | 8 | 40 | 2.5 | 0.92-6.8 |  |  |
|  | >10 | 111 | 13 | 4 | 20 | 2.5 | 0.74-8.5 |  |  |
| Condom Use | Always | 213 | 24 | 5 | 25 | Reference | | Reference | |
|  | Most of the time | 149 | 17 | 3 | 15 | 0.86 | 0.20-3.7 |  |  |
|  | Half of the time | 48 | 5.4 | 1 | 5 | 0.89 | 0.10-7.8 |  |  |
|  | Occasionally | 143 | 16 | 3 | 15 | 0.89 | 0.21-3.8 |  |  |
|  | Never | 335 | 38 | 8 | 40 | 1.0 | 0.33-3.2 |  |  |
| Sexual Transmitted | Yes | 58 | 7 | 2 | 10 | Reference | | Reference | |
| Infeccion | No | 830 | 93 | 18 | 90 | 0.63 | 0.14-2.8 |  |  |
| Parity | 0 | 664 | 75 | 14 | 70 | Reference | | Reference | |
|  | 1 | 134 | 15 | 5 | 25 | 1.8 | 0.63-5.0 |  |  |
|  | ≥ 2 | 90 | 10 | 1 | 5 | 0.53 | 0.07-4.1 |  |  |
| Smoking status | Yes | 174 | 20 | 3 | 15 | Reference | | Reference | |
|  | No | 714 | 80 | 17 | 85 | 1.4 | 0.40-4.8 |  |  |
| Cytology | NILM | 815 | 92 | 15 | 75 | Reference | | Reference | |
|  | ASCUS | 34 | 3.8 | 1 | 5 | 1.6 | 0.20-12 |  |  |
|  | LSIL | 28 | 3.2 | 3 | 15 | **5.8** | **1.6-21** | **6.3** | **1.8-24** |
|  | ASC-H/AGC_FN/HSIL | 10 | 1.1 | 1 | 5 | 5.4 | 0.65-45 |  |  |
| **Variable** |  | **n** | **Mean** | **n** | **Mean** |  |  |  |  |
| Age (years) |  | 888 | 29.4 | 20 | 29.6 | 1.1 | 0.87-1.5 |  |  |
| Age sexual debut |  | 888 | 17.6 | 20 | 16.7 | 0.83 | 0.66-1.0 |  |  |
| Years of smoking |  | 169 | 11 | 3 | 15 | 1.0 | 0.92-1.1 |  |  |

**Supplementary material table 3:** Factors associated with the detection of HPV 31

**Supplementary material table 4:** Factors associated with the detection of HPV 51

|  |  | **HPV 51** | | | | **Univariant ^a^** | | **Multivariant ^a^** | |
| --- | --- | --- | --- | --- | --- | --- | --- | --- | --- |
|  |  | **No detected** | | **detected** | |  |  |  |  |
| **Variable** | **Category** | **n** | **%** | **n** | **%** | **OR** | **95% CI** | **aOR** | **95% CI** |
| Region | La Rioja | 189 | 22 | 7 | 16 | Reference | | Reference | |
|  | Guadalajara | 287 | 33 | 15 | 33 | 1.4 | 0.56-3.5 |  |  |
|  | Basque Country | 387 | 45 | 23 | 51 | 1.6 | 0.68-3.8 |  |  |
| Vaccination | Yes | 494 | 57 | 33 | 73 | Reference | | Reference | |
|  | No | 369 | 43 | 12 | 27 | **0.49** | **0.25-0.96** | 0.90 | 0.42-1.9 |
| Gardasil vaccine | Yes | 429 | 50 | 31 | 69 | Reference | | Reference | |
|  | No | 434 | 50 | 14 | 31 | **0.45** | **0.23-0.85** | 0.59 | 0.29-1.2 |
| Vaccination before | Yes | 435 | 50 | 31 | 69 | Reference | | Reference | |
| sexual debut | No | 428 | 50 | 14 | 31 | **0.46** | **0.24-0.88** | 0.70 | 0.33-1.5 |
| Marital status | Married/Civil Partnership | 242 | 28 | 5 | 11 | Reference | | Reference | |
|  | Single | 604 | 70 | 40 | 89 | **3.2** | **1.3-8.3** | **2.4** | **0.86-6.4** |
|  | Divorced | 17 | 2 | 0 | 0 | NA | *p=* 0.29 |  |  |
| Origen | Spain | 596 | 69 | 31 | 69 | Reference | | Reference | |
|  | Latin America | 216 | 25 | 10 | 22 | 0.89 | 0.43-2.0 |  |  |
|  | Rest of the world | 51 | 6 | 4 | 9 | 2.0 | 0.51-4.4 |  |  |
| Previous screening | Yes | 691 | 80 | 31 | 69 | Reference | | Reference | |
|  | No | 172 | 20 | 14 | 31 | 1.8 | 0.94-3.5 |  |  |
| Sexual orientation | Heterosexual | 785 | 91 | 41 | 91 | Reference | | Reference | |
|  | Bisexual | 52 | 6 | 4 | 8.9 | 1.5 | 0.51-4.3 |  |  |
|  | Homosexual | 26 | 3 | 0 | 0 | NA | *p=* 0.25 |  |  |
| Sexual partners | 1-5 | 544 | 63 | 19 | 42 | Reference | | Reference | |
|  | 6-10 | 218 | 25 | 12 | 27 | 1.6 | 0.75-3.3 |  |  |
|  | >10 | 101 | 12 | 14 | 31 | **4.0** | **1.9-8.2** | **2.6** | **1.2-5.6** |
| Condom Use | Always | 205 | 24 | 13 | 29 | Reference | | Reference | |
|  | Most of the time | 141 | 16 | 11 | 24 | 1.2 | 0.54-2.8 |  |  |
|  | Half of the time | 44 | 5 | 5 | 11 | 1.8 | 0.60-5.3 |  |  |
|  | Occasionally | 142 | 16 | 4 | 9 | 0.44 | 0.14-1.4 |  |  |
|  | Never | 331 | 38 | 12 | 27 | 0.57 | 0.26-1.3 |  |  |
| Sexual Transmitted | Yes | 55 | 6.4 | 5 | 11 | Reference | | Reference | |
| Infection | No | 808 | 94 | 40 | 89 | 0.54 | 0.21-1.4 |  |  |
| Parity | 0 | 641 | 74 | 37 | 82 | Reference | | Reference | |
|  | 1 | 133 | 15 | 6 | 13 | 0.78 | 0.32-1.9 |  |  |
|  | ≥ 2 | 89 | 10 | 2 | 4 | 0.39 | 0.09-1.7 |  |  |
| Smoking status | Yes | 161 | 19 | 16 | 36 | Reference | | Reference | |
|  | No | 702 | 81 | 29 | 64 | **0.42** | **0.22-0.79** | 0.55 | 0.27-1.1 |
| Cytology | NILM | 799 | 93 | 31 | 70 | Reference | | Reference | |
|  | ASCUS | 32 | 4 | 3 | 7 | 2.4 | 0.70-8.3 |  |  |
|  | LSIL | 22 | 3 | 9 | 21 | **11** | **4.4-25** | **8.2** | **3.3-21** |
|  | ASC-H/AGC_FN/HSIL | 10 | 1.2 | 1 | 2.3 | 2.6 | 0.32-21 |  |  |
| **Variable** |  | **n** | **Mean** | **n** | **Mean** |  |  |  |  |
| Age (years) |  | 863 | 29.4 | 45 | 28.7 | 0.76 | **0.63-0.93** | 0.82 | 0.66-1.0 |
| Age sexual debut |  | 863 | 17.6 | 45 | 17.2 | 0.93 | 0.82-1.1 |  |  |
| Years of smoking |  | 157 | 11.3 | 15 | 8.8 | 1.04 | 0.98-1.1 |  |  |

a: statistically significant results are shown in bold. NA: Not Applicable, p value provided.

**Supplementary material table 5:** Factors associated with the detection of HPV 70

|  |  | **HPV 70** | | | | **Univariant** | | **Multivariant** | |
| --- | --- | --- | --- | --- | --- | --- | --- | --- | --- |
|  |  | **No detected** | | **detected** | |  |  |  |  |
| **Variable** | **Category** | **n** | **%** | **n** | **%** | **OR** | **95% CI** | **aOR** | **95% CI** |
| Region | La Rioja | 191 | 21 | 5 | 29 | Reference | | Reference | |
|  | Guadalajara | 298 | 33 | 4 | 24 | 0.51 | 0.14-2.0 |  |  |
|  | Basque Country | 402 | 45 | 8 | 47 | 0.76 | 0.25-2.4 |  |  |
| Vaccination | Yes | 513 | 58 | 14 | 82 | Reference | | Reference | |
|  | No | 378 | 42 | 3 | 18 | **0.29** | **0.08-1.0** | 0.28 | 0.07-1.0 |
| Gardasil vaccine | Yes | 449 | 50 | 11 | 65 | Reference | | Reference | |
|  | No | 442 | 50 | 6 | 35 | 0.55 | 0.20-1.5 |  |  |
| Vaccination before | Yes | 454 | 51 | 12 | 71 | Reference | | Reference | |
| Sexual debut | No | 437 | 49 | 5 | 29 | 0.43 | 0.15-1.2 |  |  |
| Marital status | Married/Civil Partnership | 242 | 27 | 5 | 29 | Reference | | Reference | |
|  | Single | 632 | 71 | 12 | 71 | 0.92 | 0.32-2.6 |  |  |
|  | Divorced | 17 | 2 | 0 | 0 | NA | *p=* 0.57 |  |  |
| Origen | Spain | 615 | 69 | 12 | 71 | Reference | | Reference | |
|  | Latin America | 223 | 25 | 3 | 18 | 0.69 | 0.19-2.5 |  |  |
|  | Rest of the world | 53 | 6 | 2 | 12 | 1.9 | 0.42-8.9 |  |  |
| Previous screening | Yes | 708 | 79 | 14 | 82 | Reference | | Reference | |
|  | No | 183 | 21 | 3 | 18 | 0.83 | 0.24-3.0 |  |  |
| Sexual orientation | Heterosexual | 810 | 91 | 16 | 94 | Reference | | Reference | |
|  | Bisexual | 55 | 6.2 | 1 | 5.9 | 0.92 | 0.12-7.1 |  |  |
|  | Homosexual | 26 | 3 | 0 | 0 | NA | *P=* 0.47 |  |  |
| Sexual partners | 1-5 | 557 | 63 | 6 | 35 | Reference | | Reference | |
|  | 6-10 | 222 | 25 | 8 | 47 | **3.3** | **1.1-9.8** | 2.7 | 0.89-8.1 |
|  | >10 | 112 | 13 | 3 | 18 | 2.5 | 0.61-10 |  |  |
| Condom Use | Always | 216 | 24 | 2 | 12 | Reference | | Reference | |
|  | Most of the time | 146 | 16 | 6 | 35 | 4.4 | 0.87-23 |  |  |
|  | Half of the time | 49 | 5.5 | 0 | 0 | NA | *p=* 0.31 |  |  |
|  | Occasionally | 144 | 16 | 2 | 12 | 1.5 | 0.21-11 |  |  |
|  | Never | 336 | 38 | 7 | 41 | 2.3 | 0.46-11 |  |  |
| Sexual Transmitted | Yes | 58 | 6.5 | 2 | 12 | Reference | | Reference | |
| Infection | No | 833 | 93 | 15 | 88 | 0.52 | 0.12-2.3 |  |  |
| Parity | 0 | 663 | 74 | 15 | 88 | Reference | | Reference | |
|  | 1 | 137 | 15 | 2 | 12 | 0.65 | 0.15-2.9 |  |  |
|  | ≥ 2 | 91 | 10 | 0 | 0 | NA | *p=* 0.15 |  |  |
| Smoking status | Yes | 169 | 19 | 8 | 47 | Reference | | Reference | |
|  | No | 722 | 81 | 9 | 53 | **0.26** | **0.10-0.70** | **0.36** | **0.13-0.99** |
| Cytology | NILM | 817 | 92 | 13 | 76 | Reference | | Reference | |
|  | ASCUS | 35 | 3.9 | 0 | 0 | NA | *p=* 0.46 |  |  |
|  | LSIL | 28 | 3.2 | 3 | 18 | **6.7** | **1.8-25** | **5.0** | **1.3-20** |
|  | ASC-H/AGC_FN/HSIL | 10 | 1.1 | 1 | 5.9 | 6.3 | 0.74-53 |  |  |
| **Variable** |  | **n** | **Mean** | **n** | **Mean** |  |  |  |  |
| Age (years) |  | 891 | 29.4 | 17 | 29.1 | 0.88 | 0.65-1.2 |  |  |
| Age sexual debut |  | 891 | 17.6 | 17 | 16.9 | 0.89 | 0.71-1.1 |  |  |
| Years of smoking |  | 164 | 11.2 | 8 | 8.8 | 0.89 | 0.76-1.0 |  |  |

a: statistically significant results are shown in bold. NA: Not Applicable, p value provided

**Questionnaire**

**EPIDEMIOLOGICAL DATA**

**Study region:**

☐ Guadalajara	☐ La Rioja ☐ Basque Country
**Age:** ____

**Marital status:**

☐ Married/Civil Partnership	☐ Single	☐ Divorced/Separated	☐ Widowed

**Region of origin:**

☐ Spain ☐ Latin America ☐ Africa ☐ Europe ☐ Others: ______

**SCREENING AND VACCINATION DATA**

**Previous cervical cancer screening test in public or private healthcare:**

☐ No	☐ Yes

**Vaccinated against HPV^1,2^:**

☐ No	☐ Yes

**Fully vaccinated^3^:**

☐ No	☐ Yes

**Vaccinated into the regional Immunization program (schools/health center):**

☐ No	☐ Yes

**Vaccination Place^4^:**

Spanish Autonomous Community: _____

Out of Spain ☐

**Vaccine type:**

☐ Cervarix ☐ Gardasil ☐ Gardasil 9 ☐ Unknown

**HPV vaccinated before first sexual intercourse:**

☐ No	☐ Yes

**SEXUAL BEHAVIOR DATA**

**Age at first sexual intercourse:** ____

**Sexual orientation:**

☐ Heterosexual	☐ Homosexual	☐ Bisexual

**Number of sexual partners in lifetime:**

☐ 0–5	☐ 6–10	☐ 11–20	☐ 21–50	☐ More than 50

**Condom use during sexual intercourse in the past year:**

☐ Never	☐ Occasionally	☐ About half the time

☐ Most of the time	☐ Always

**Sexual transmitted infections in lifetime:** ☐ No	☐ Yes

☐ Chlamydia infection ☐ Genital herpes	☐ Gonorrhea

☐ HIV infection	☐ Genital warts/Condyloma

☐ Trichomoniasis	☐ Syphilis	☐ Others: ______

**OTHER DATA**

**Parity:** ☐ 0	☐ 1	☐ ≥ 2

**Current smoker:** ☐ No ☐ Yes	**Number of years smoking:** ______

**1.** HPV vaccination data were collected from medical records and information provided by the participant.

**2.** Definition: Participants who had received at least one dose of any HPV vaccine.

**3.** Vaccinated with 2 o three doses according to the age.

**4.** Identifying the Spanish region in which the participant was vaccinated allows subsequent determination of the vaccine type based on data available from each regional immunization program.
